# Estimates and determinants of health facility delivery in the Birhan cohort in Ethiopia

**DOI:** 10.1101/2023.08.04.23293667

**Authors:** Bezawit M. Hunegnaw, Frederick G. B. Goddard, Delayehu Bekele, Sebastien Haneuse, Clara Pons-Duran, Mesfin Zeleke, Yahya Mohammed, Chalachew Bekele, Grace J. Chan

## Abstract

Health facility delivery is one of the critical indicators to monitor progress towards the provision of skilled delivery care and reduction in perinatal mortality. In Ethiopia, utilization of health facilities for skilled delivery care has been increasing but varies greatly by region and among specific socio-demography groups. We aim to measure the prevalence and determinants of health facility delivery in the Amhara region in Ethiopia.

From December 2018 to November 2020, we conducted a longitudinal study from a cohort of 2801 pregnant women and described the location of delivery and the association with determinants. We interviewed a subset of women who delivered in the community and analyzed responses using the three delays model to understand reasons for not using health facility services. A multivariable poisson regression model with robust error variance was used to estimate the presence and magnitude of association between location of delivery and the determinants.

Of the 2,482 pregnant women followed through to birth, 73.6% (n=1,826) gave birth in health facilities, 24.3% (n=604) gave birth at home and 2.1% (n=52) delivered on the way to a health facility. Determinants associated with increased likelihood of delivery at a health facility included formal maternal education, shorter travel times to health facilities, primiparity, higher wealth index and having attended at least one ANC visit. Most common reasons mothers gave for not delivering in a health facility were delays in individual/family decision to seek care. The proportion of deliveries occurring in health facilities is improving but falls below targets. Interventions that focus on the identified social-demographic determinants to improve uptake of health facility delivery care are warranted.

## Introduction

Globally, an estimated 810 maternal deaths and 12,000 perinatal deaths occur every day, mostly from preventable causes related to pregnancy and childbirth (1–3). Most of these deaths occur disproportionately in low- and middle-income countries. Sub-Saharan Africa in particular continues to suffer the highest rates, accounting for nearly two-thirds of all maternal deaths and two-fifths of global newborn deaths (4,5). Ethiopia is one of the countries in this category where maternal and perinatal mortality rates continue to be high.

Effective interventions and care packages known to prevent maternal and perinatal mortality include emergency obstetric and newborn care, active management of third stage of labor, blood transfusion, management of sepsis, all of which are provided in the setup of health facilities. Delivery in a health facility with the provision of skilled care during and immediately after delivery is a key strategy that can improve perinatal outcomes by providing clinical assessment to detect and manage complications, provide timely and lifesaving interventions, as well as referrals, when needed (6–8). It is estimated that 54% of maternal deaths and 43% of neonatal deaths can be prevented each year with the provision of skilled care during delivery (9,10). However, many women give birth outside of health facilities with no access to such care and interventions, and are therefore at risk for serious complications, such as hemorrhage or sepsis (11).

The Sustainable Development Goal 3 (SDG 3) has set a target for all countries to lower the Maternal Mortality Ratio (MMR) to less than 70 per 100,000 of live births by 2030. The proportion of births attended by skilled birth attendants (SBAs) is one of the critical progress indicators used to monitor the achievement of this goal to reduce maternal and newborn morbidity and mortality (11–13). Under SDG 3.1, the aim is to increase the proportion of births assisted by SBAs to 90% (14). In order to achieve this in Ethiopia, the national Health Sector Transformation Plan II (HSTP II) aims to increase the rate of institutional delivery to 76% by 2024-25, hence increasing the proportion of deliveries with access to skilled delivery care and interventions.

According to the Ethiopian Demographic Health Survey (EDHS), the proportion of deliveries occurring in health facilities have increased from 10% in 2011 to 26% in 2016 and 48% in 2019 (15,16). However, the figures vary widely across and within the country’s regions, with 95% of births delivered in health facilities in the capital city Addis Ababa in contrast to 28% in the Afar region (16). The most recent estimates from 2019 report that four of the eleven regions in Ethiopia have health facility delivery rates below 50%, five of the regions (including the Amhara region) reported skilled birth attendance between 50% and 70%, and the remaining two regions had 70% or more deliveries in health facilities (16). This underlines the wide gaps in utilization of facility-based delivery services in different parts of Ethiopia that are far from national and global targets.

Several factors at individual, household and community levels have been identified as key determinants that influence the location of delivery. In different contexts in sub-Saharan Africa, factors such as place of residence, household wealth, education and distance to health facilities have been shown to determine where women give birth (17–22). In the Ethiopian context, there have been observations of significant heterogeneity based on individual and community level factors showing varying degrees of association of these determinants to location of delivery (23–28). Trends of health facility delivery have been dynamic, with most regions showing improvement in utilization of facility-based skilled delivery care services (15,16). However, some regions have shown a decline (29). Therefore, it is important to identify the magnitude and direction of change in the utilization of these key services, as well as understand the effect of determinants on different populations and subgroups to identify inequalities and ensure equitable distribution of service delivery. The evidence generated from this study will be useful to understand progress, model successes and take best practices to scale as well as recommend key interventions to increase the prevalence of skilled delivery attendance. We aim to provide estimates of health facility deliveries and describe the factors that affect location of delivery in the Amhara region of Ethiopia from 2018 to 2020.

## Methods

### Study design and study setting

We conducted a pregnancy and birth cohort study, the Birhan maternal and child health (MCH) cohort nested within the Birhan field site (30). The field site is in the North Shewa zone of Ethiopia, located 130 km north of the capital city, Addis Ababa (31). The catchment area consists of two districts, Angolela Tera and Kewet/Shewa Robit, covering a population of 79,653 individuals and 19,957 households. Each district is composed of eight clusters (kebeles). The area is predominantly rural.

### Study population

Pregnancies within the HDSS were identified through pregnancy screening during quarterly house to house surveillance visits within the study catchment area or during visits to health facility (30). The Birhan MCH cohort study enrolls consenting pregnant women within the catchment area and follows them for birth and birth outcomes. Pregnancy outcomes were ascertained through scheduled follow up of the cohort at home and in the health facilities. Our analysis included data collected from all pregnant women followed through to birth from December 2018 to November 2020 (30). We excluded data if pregnancies were not followed through to birth, for example due to miscarriage, death, outmigration, or lost to follow-up.

### Definition of outcomes and determinants

The primary outcome for this study was location of delivery, and whether this was in a health facility or in the community. Facility/institutional delivery was defined as delivery occurring in a hospital, primary level health center or a clinic. Community delivery was defined as delivery occurring outside of a health institution, either at home or on the way to a health institution.

To assess wealth, we used principal component analysis of household assets to categorize households into five distinct quintiles according to the guidelines of the DHS program (DHS program). The living conditions and assets included in the PCA to create the index were selected based on the list of variables used in the Ethiopian DHS 2019. Parity was defined as the number of times a woman has given birth and responses were coded as primiparous for women who gave birth for the first time and multiparous for those who had more than one birth. Maternal formal education was categorized into three levels to include those with no formal education, those with only primary level education and those with secondary level of education or higher. To estimate the effect of distance from mothers’ residence to closest health facility, we used maternal estimates of travel times to the health facility.

### Data collection

The Birhan field site includes quarterly house-to-house surveillance and data collection on household sociodemographic and baseline individual information as well as pregnancy screening and identification. Follow up data for members of the MCH cohort were collected through scheduled home visits and facility visits when women presented for antenatal care or delivery. Scheduled home visits were conducted every three months until 32 weeks of gestation, every two weeks until 36 weeks of gestation and subsequently, every week until delivery. To assist in identification of community births, several birth notification mechanisms were employed, including key community informants and self-notification by families. A structured Open Data Kit based electronic data collection system was used to enter responses and observational data that were then sent to a central, secured server. Participant data were deidentified and authors had no access to information that could identify participants during or after data collection.

To collect data on the main outcome (location of delivery), home-based interviews and facility-based study observations were performed. Responses were categorized as births that occurred in a health facility, at home, or on the way to a health facility. For the purposes of further analyses, women who delivered at home and on the way to a health facility were grouped together as community deliveries, given that only few women delivered on the way to a health facility to be considered as a separate group for analysis. Additionally, it may be assumed that women who delivered on the way to a facility may have attempted to deliver at home, but later decided to deliver in a health facility.

To describe association of determinants to location of delivery, covariates were selected based on literature review which included sociodemographic characteristics such as place of residence (rural vs urban), wealth index, level of education, maternal age, distance to health facility, ethnicity, and religion as well as obstetric characteristics such as parity, number of ANC visits and day of delivery. These were further categorized into four themes: socio-cultural factors, perceived benefit/need of skilled attendance, economic accessibility, and physical accessibility (32). To understand the reasons for home delivery, women who were reached within the first week of birth were asked about their main reason for home delivery.

### Statistical analysis

Descriptive analyses were performed to estimate the prevalence of institutional deliveries as well as the differences in characteristics between women who delivered in the different locations. To describe presence and magnitude of associations between determinants and outcome, a multivariable poisson regression analysis with robust error variance was performed, presented as follows:

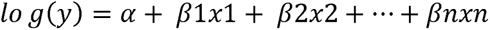

Where,

Y is the dependent variable,

β is the regression coefficient,

x is the predictor variable

Considering the main outcome to be more common (more than 10%), we measured associations between the factors and outcome using relative risk instead of odds ratios in order to avoid overestimation of the relationship. Bivariate associations between each covariate and location of delivery were explored independently. Variables that were statistically significant at p < 0.05 were entered together into the multivariable model. In the multivariable model, those considered not statistically significant at p <0.05 were removed from the model and re-entered one at a time, keeping only those that became statistically significant. Presence of collinearity was assessed by computing the variance inflation factor and adjusting for any highly correlated variables. To describe any variability of the outcome based on community clusters, a random effect regression model was employed. Extraction of dependent and independent variables from the longitudinal dataset and preliminary analyses were done using R Version 4.0.2 and Stata Version 15.1.

Among women who delivered in the community, we described the mothers’ reasons for not delivering in a health facility. Data from coded responses and free text were summarized in thematic areas and presented.

To consider the possible effect of loss to follow up on our findings, we performed sensitivity analyses to re-estimate prevalence assuming all pregnancies lost to follow-up resulted in community births or facility births.

### Ethics statement

This study was approved by the Institutional Review Boards (IRBs) of St Paul’s Hospital Millennium Medical College (SPHMMC), Addis Ababa, Ethiopia, under approval number [PM 23/274] and Boston Children’s Hospital, Boston, MA, United States, under approval number [IRB-P00028224]. Formal informed consent was obtained from eligible pregnant women prior to enrollment into the cohort.

## Results

A total of 2,801 pregnant women were identified and enrolled to the study. Pregnancy outcomes follow up was completed for 2,628 (93.8%) participants. The overall cohort characteristics and birth outcome information are published elsewhere (33). Among those with complete pregnancy outcome follow up, 2,482 women, whose pregnancies were in the gestational age range of 28 to 46 weeks at birth, were included in this analysis (Table 1)

**Table 1:**
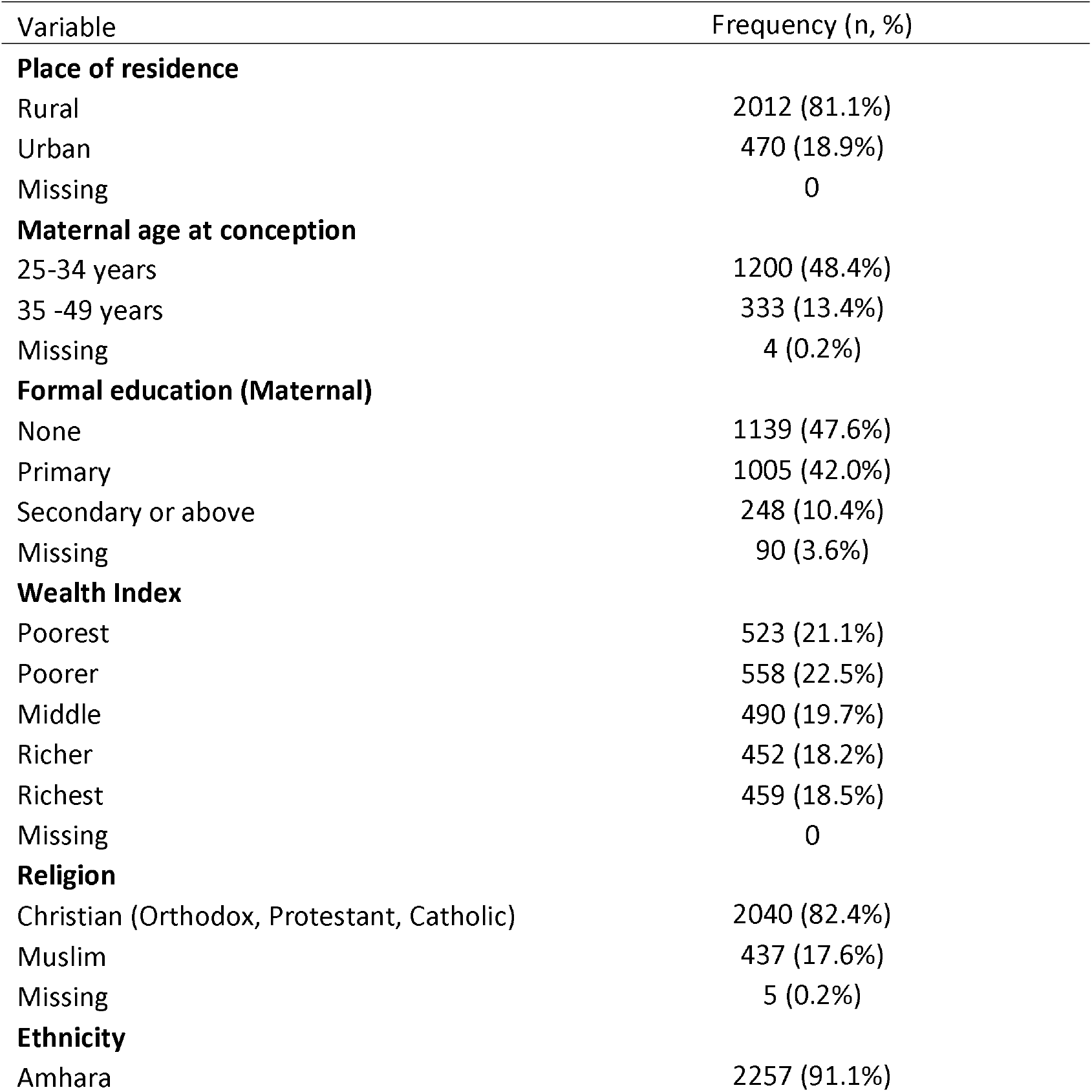
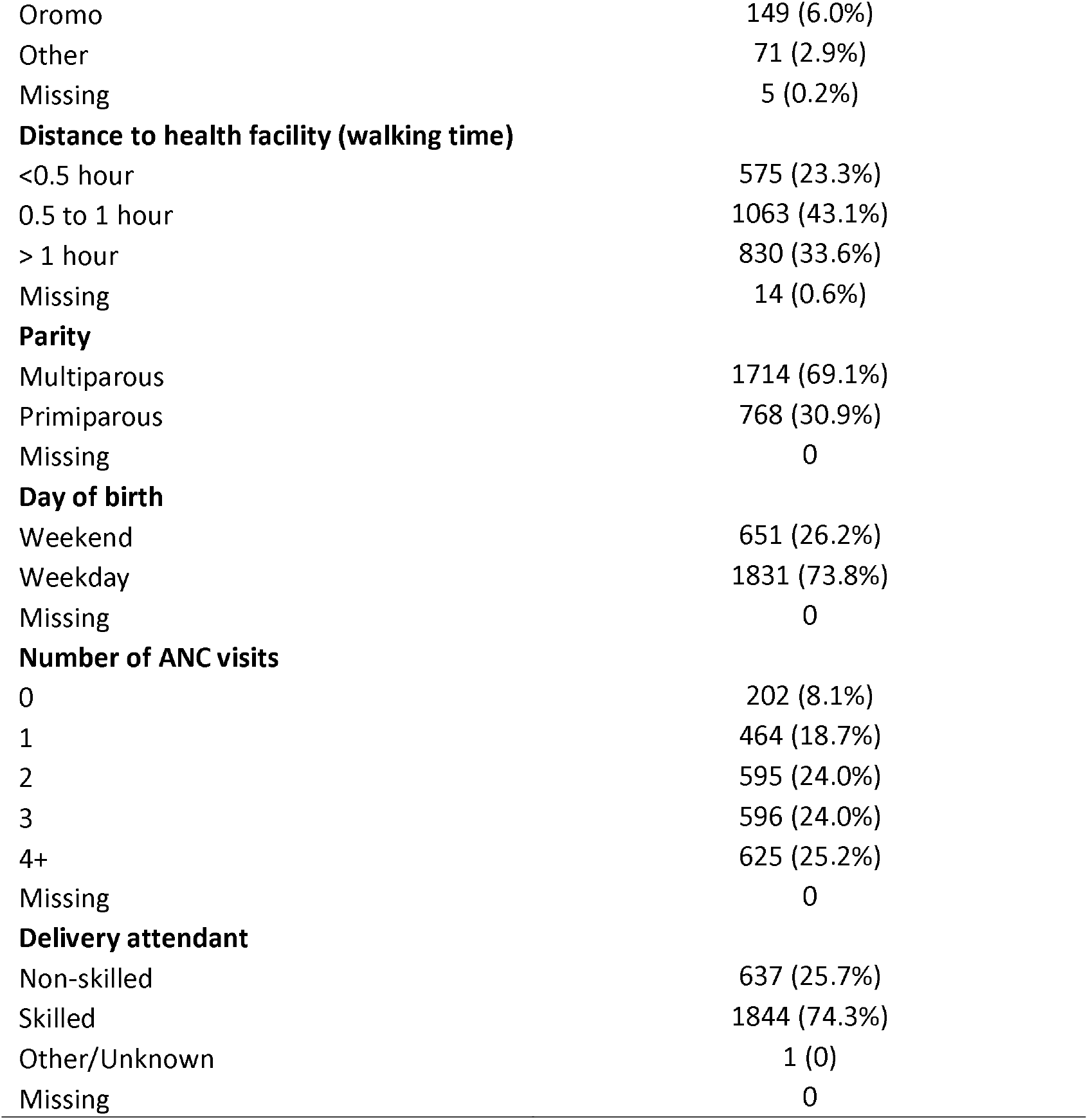
Sociodemographic and Obstetric characteristics of women enrolled in the Birhan MCH cohort, 2018-2020 (n=2482)

| Variable | Frequency (n, %) |
| --- | --- |
| **Place of residence** |  |
| Rural | 2012 (81.1%) |
| Urban | 470 (18.9%) |
| Missing | 0 |
| **Maternal age at conception** |  |
| 25-34 years | 1200 (48.4%) |
| 35 -49 years | 333 (13.4%) |
| Missing | 4 (0.2%) |
| **Formal education (Maternal)** |  |
| None | 1139 (47.6%) |
| Primary | 1005 (42.0%) |
| Secondary or above | 248 (10.4%) |
| Missing | 90 (3.6%) |
| **Wealth Index** |  |
| Poorest | 523 (21.1%) |
| Poorer | 558 (22.5%) |
| Middle | 490 (19.7%) |
| Richer | 452 (18.2%) |
| Richest | 459 (18.5%) |
| Missing | 0 |
| **Religion** |  |
| Christian (Orthodox, Protestant, Catholic) | 2040 (82.4%) |
| Muslim | 437 (17.6%) |
| Missing | 5 (0.2%) |
| **Ethnicity** |  |
| Amhara | 2257 (91.1%) |
| Oromo | 149 (6.0%) |
| Other | 71 (2.9%) |
| Missing | 5 (0.2%) |
| **Distance to health facility (walking time)** |  |
| <0.5 hour | 575 (23.3%) |
| 0.5 to 1 hour | 1063 (43.1%) |
| > 1 hour | 830 (33.6%) |
| Missing | 14 (0.6%) |
| **Parity** |  |
| Multiparous | 1714 (69.1%) |
| Primiparous | 768 (30.9%) |
| Missing | 0 |
| **Day of birth** |  |
| Weekend | 651 (26.2%) |
| Weekday | 1831 (73.8%) |
| Missing | 0 |
| **Number of ANC visits** |  |
| 0 | 202 (8.1%) |
| 1 | 464 (18.7%) |
| 2 | 595 (24.0%) |
| 3 | 596 (24.0%) |
| 4+ | 625 (25.2%) |
| Missing | 0 |
| **Delivery attendant** |  |
| Non-skilled | 637 (25.7%) |
| Skilled | 1844 (74.3%) |
| Other/Unknown | 1 (0) |
| Missing | 0 |

The mean age of the participants at enrollment was 27 years (± 6 years). Most women (48.4%) were between 25 to 34 years of age, while 38.1% were between ages 15 and 24. Of the women in the cohort, 81.1% lived in rural areas and 43.6% came from poor households in the lowest two quintiles. No formal education was reported by 47.6% of the participants, while 42% had some primary level education. The Amhara ethnic group made up 91.1% of the study participants with 6% from the Oromo ethnic group. Religious groups were identified as Christians (82.4%) and Muslims (17.6%). Multiparous women accounted for 69% of the study population. During their pregnancy, 91.9% of women had at least one ANC visit, 25.2% had 4 or more ANC visits, while 8.1% had no recorded or reported ANC visits. (Table 1)

### Proportion of births in health facilities

During the study period, out of a total of 2,482 women, 1,826 women (73.6%) gave birth in health facilities, 604 births (24.3%) occurred at home and 52 women (2.1%) delivered on the way to a health facility. All of the births in health facilities and 42.3% (n=22) of the births that occurred on the way to a health facility received care from an SBA. None of the deliveries that occurred at home were attended by an SBA. (Table 2)

**Table 2:** Location of delivery and skilled delivery care during birth for women enrolled in the Birhan MCH cohort, 2018-2020.

| Variable | N | n (%) |
| --- | --- | --- |
| **Place of delivery** |  |  |
| Health facility | 2482 | 1826 (73.6%) |
| Home | 2482 | 604 (24.3%) |
| On the way to health facility | 2482 | 52 (2.1%) |
| **Delivered by SBA** |  |  |
| Health facility | 1826 | 1822 (99.8%) |
| Home | 604 | 0 (0) |
| On the way to health facility | 52 | 22 (42.3%) |

### Bivariate analysis

Women living in urban areas (RR = 1.29; 95% CI 1.23-1.34), having primary level education (RR = 1.22; 95% CI 1.16-1.29), secondary education or above (RR = 1.48; 95% CI 1.40-1.56), and primiparous (RR = 1.31; 95% CI 1.25-1.37) had a higher likelihood of delivering in a health facility. Having longer travel times to the nearest health facility of half an hour to an hour (RR = 0.82; 95% CI 0.79-0.86), and over an hour (RR = 0.62; 95% CI 0.58-0.66) reduced the likelihood of institutional delivery. Women between 35– 49 years old were less likely to deliver in a health facility compared to their younger counterparts (RR = 0.82; 95% CI 0.76-0.90). Differences in location of delivery were also observed among different religions; Muslim women were less likely to deliver in a health facility (RR = 0.88; 95% CI 0.82-0.94) compared to women from Christian religion. Women who had at least one ANC visit were more likely to deliver in a health facility, with women who had 4 or more ANC visits having the highest likelihood of facility delivery (RR = 1.78; 95% CI 1.54-2.06). Household wealth was also associated with place of delivery, with women from wealthier households having increased likelihood to deliver in a health facility. No statistically significant differences were observed from the bivariate analysis on location of delivery based on days of the week (weekday vs weekend) when the women delivered (Table 3).

**Table 3:**
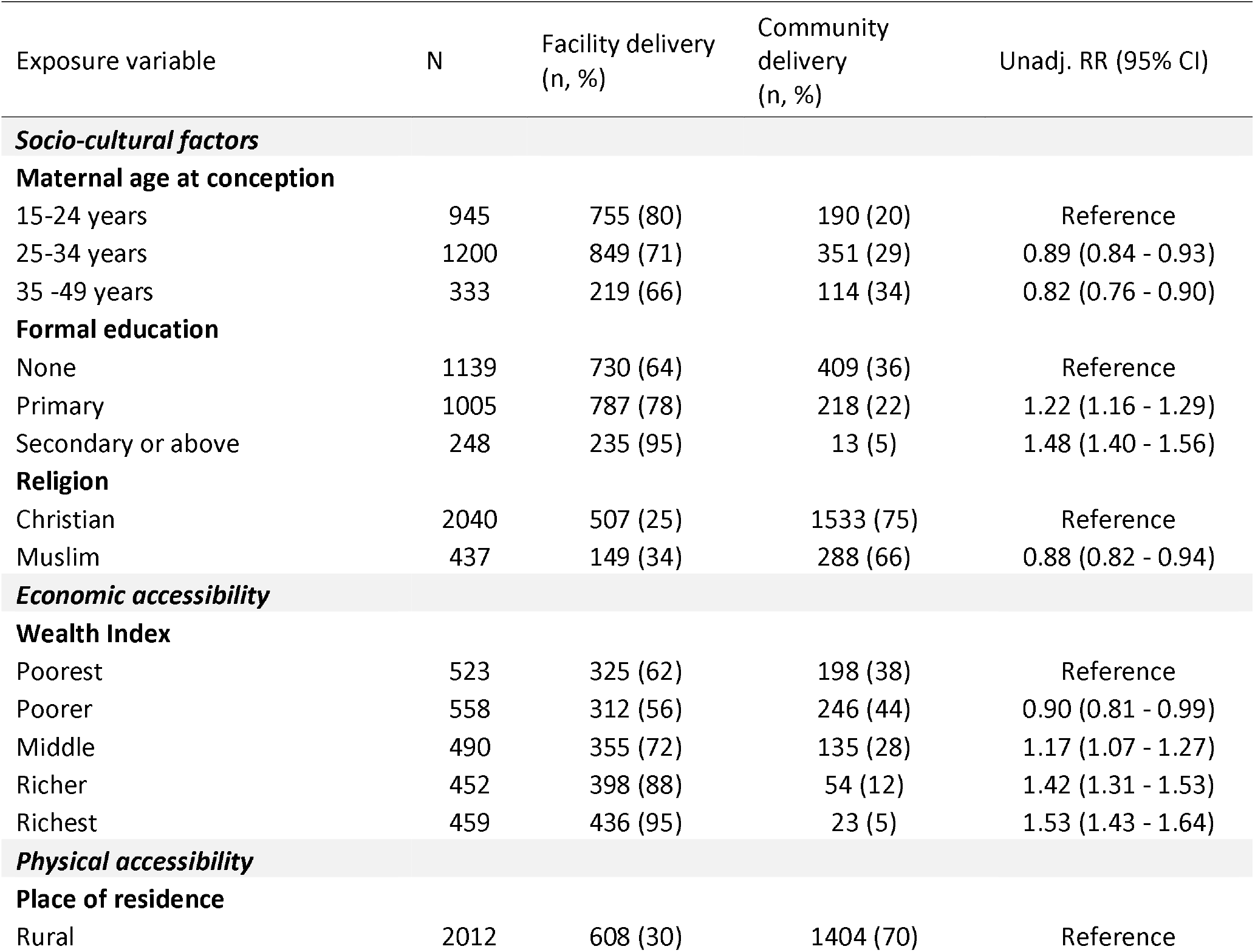
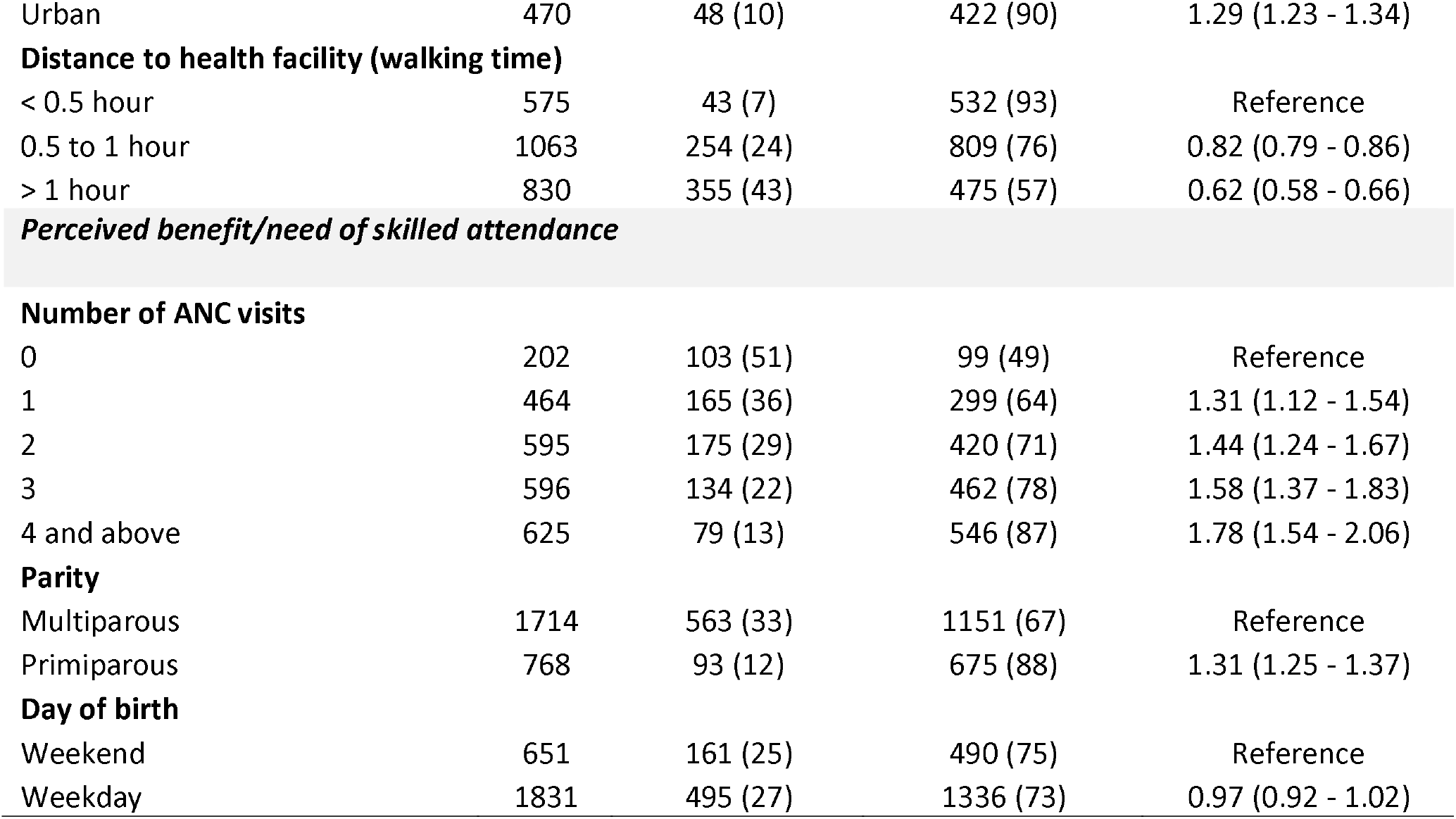
Unadjusted relative risk ratio for determinants of facility delivery for women enrolled in the Birhan MCH cohort, 2018-2020.

| Exposure variable | N | Facility delivery<br>(n, %) | Community<br>delivery<br>(n, %) | Unadj. RR (95% CI) |
| --- | --- | --- | --- | --- |
| ***Socio-cultural factors*** |  |  |  |  |
| **Maternal age at conception** |  |  |  |  |
| 15-24 years | 945 | 755 (80) | 190 (20) | Reference |
| 25-34 years | 1200 | 849 (71) | 351 (29) | 0.89 (0.84 - 0.93) |
| 35 -49 years | 333 | 219 (66) | 114 (34) | 0.82 (0.76 - 0.90) |
| **Formal education** |  |  |  |  |
| None | 1139 | 730 (64) | 409 (36) | Reference |
| Primary | 1005 | 787 (78) | 218 (22) | 1.22 (1.16 - 1.29) |
| Secondary or above | 248 | 235 (95) | 13 (5) | 1.48 (1.40 - 1.56) |
| **Religion** |  |  |  |  |
| Christian | 2040 | 507 (25) | 1533 (75) | Reference |
| Muslim | 437 | 149 (34) | 288 (66) | 0.88 (0.82 - 0.94) |
| ***Economic accessibility*** |  |  |  |  |
| **Wealth Index** |  |  |  |  |
| Poorest | 523 | 325 (62) | 198 (38) | Reference |
| Poorer | 558 | 312 (56) | 246 (44) | 0.90 (0.81 - 0.99) |
| Middle | 490 | 355 (72) | 135 (28) | 1.17 (1.07 - 1.27) |
| Richer | 452 | 398 (88) | 54 (12) | 1.42 (1.31 - 1.53) |
| Richest | 459 | 436 (95) | 23 (5) | 1.53 (1.43 - 1.64) |
| ***Physical accessibility*** |  |  |  |  |
| **Place of residence** |  |  |  |  |
| Rural | 2012 | 608 (30) | 1404 (70) | Reference |
| Urban | 470 | 48 (10) | 422 (90) | 1.29 (1.23 - 1.34) |
| **Distance to health facility (walking time)** |  |  |  |  |
| < 0.5 hour | 575 | 43 (7) | 532 (93) | Reference |
| 0.5 to 1 hour | 1063 | 254 (24) | 809 (76) | 0.82 (0.79 - 0.86) |
| > 1 hour | 830 | 355 (43) | 475 (57) | 0.62 (0.58 - 0.66) |
| ***Perceived benefit/need of skilled attendance*** |  |  |  |  |
| **Number of ANC visits** |  |  |  |  |
| 0 | 202 | 103 (51) | 99 (49) | Reference |
| 1 | 464 | 165 (36) | 299 (64) | 1.31 (1.12 - 1.54) |
| 2 | 595 | 175 (29) | 420 (71) | 1.44 (1.24 - 1.67) |
| 3 | 596 | 134 (22) | 462 (78) | 1.58 (1.37 - 1.83) |
| 4 and above | 625 | 79 (13) | 546 (87) | 1.78 (1.54 - 2.06) |
| **Parity** |  |  |  |  |
| Multiparous | 1714 | 563 (33) | 1151 (67) | Reference |
| Primiparous | 768 | 93 (12) | 675 (88) | 1.31 (1.25 - 1.37) |
| **Day of birth** |  |  |  |  |
| Weekend | 651 | 161 (25) | 490 (75) | Reference |
| Weekday | 1831 | 495 (27) | 1336 (73) | 0.97 (0.92 - 1.02) |

### Multivariate analysis

In multivariate analysis, maternal formal education, religion, wealth index, distance to health facility, number of ANC visits and parity remained statistically significant. The influence of maternal formal education was attenuated in the multivariate model and showed marginally increased likelihood of women with primary level of education to deliver in health facilities (aRR = 1.08; 95% CI 1.02-1.13).Women from the richest households were 22% more likely to give birth in a health facility (aRR = 1.22; 95% CI 1.12 - 1.32). Similarly, ANC attendance continued to show strong positive association with location of delivery and women who attended 4 or more ANC visits had 48% increased likelihood of delivering at a health facility (aRR = 1.48; 95% CI 1.29 - 1.70). Primiparous women were also more likely to have institutional delivery than multiparous counterparts (aRR = 1.18; 95% CI 1.13 - 1.23). Conversely, women from muslim households were 20% less likely to deliver in health facilities (aRR = 0.8, 95% CI 0.74 - 0.87). Women who had more than an hour walking time to a health facility were 21% less likely to deliver in a health facility (aRR = 0.79; 95% CI 0.73 - 0.85) The effect of place of residence (rural/urban) and maternal age were not statistically significant after controlling for other variables included in the model (Table 4).

**Table 4:**
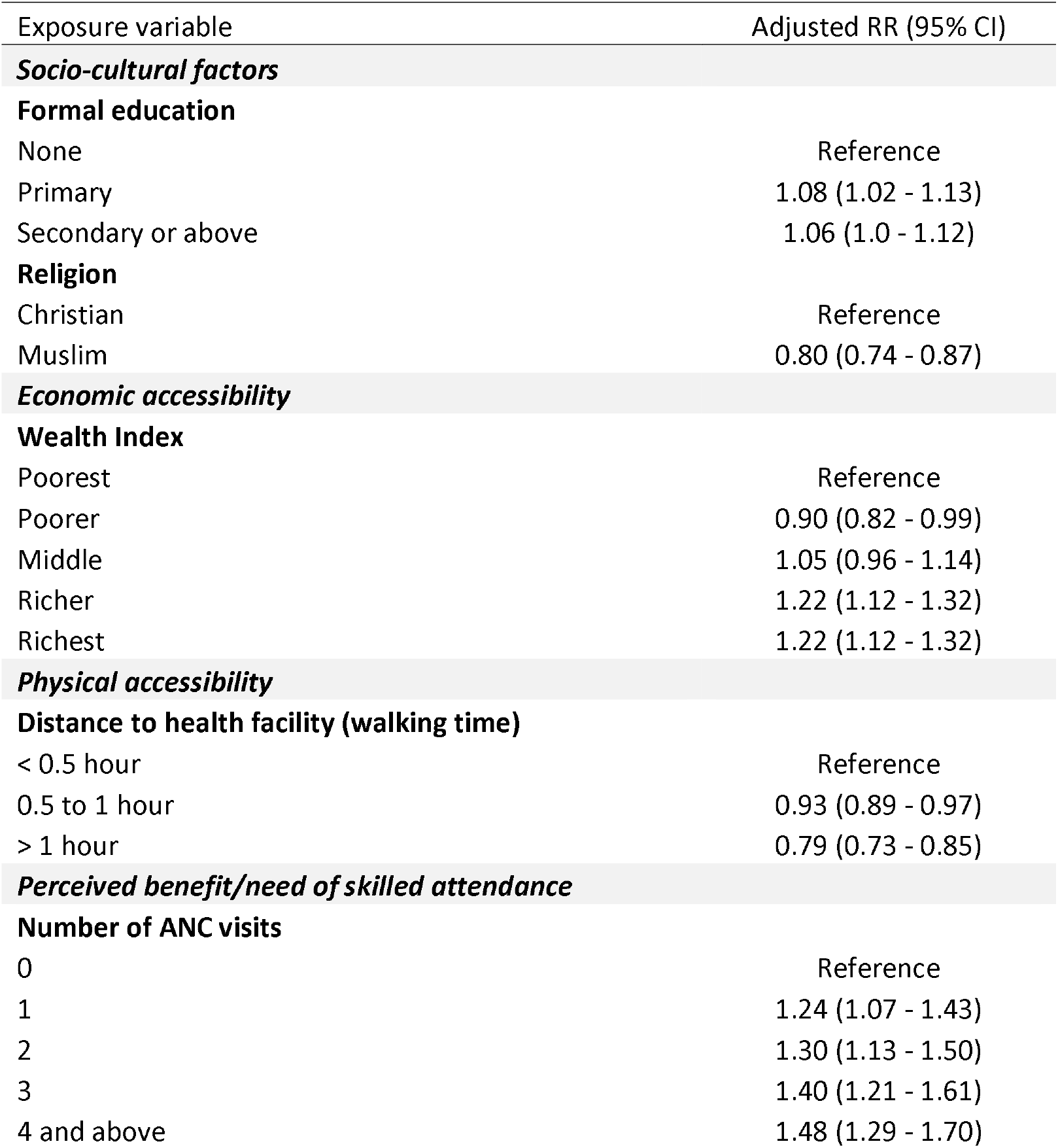
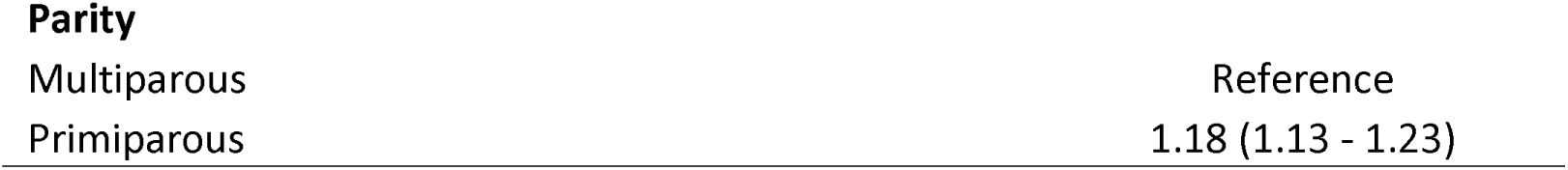
Adjusted relative risk ratio for determinants of facility delivery for women enrolled in the Birhan MCH cohort, 2018-2020.

In the null model, 13.3% of the total variance in the odds of delivering in a health facility was accounted by between cluster (*kebele*) variability (ICC 0.133; SE 0.04). In the final adjusted model, the variability remained significant, but declined to 6.4% (ICC 0.064; SE 0.03).

### Reasons for birth in the community

The specific reasons of a subset of 470 women who delivered at home were classified into three phases under the three delays model: delay in deciding to seek care (Phase I delay), delay in reaching the health facility (Phase II delay) and delay in receiving care at the health facility (Phase III delay) (Fig 1). Approximately 80% of the reasons were related to phase I delay, with the other participants referring to phase II delays. Among those with phase I delay, the most common reasons cited included perception of fast progress of labor (68.1%), maternal preference to deliver at home (23.1%) and lack of anyone to accompany the mother (7.7%). Reasons resulting in phase II delay included the health facility being too far away (85.1%) and difficulties accessing transportation, which included delayed or unreachable ambulance services (14.9%) (Fig 1 and 2).

**Fig 1:**
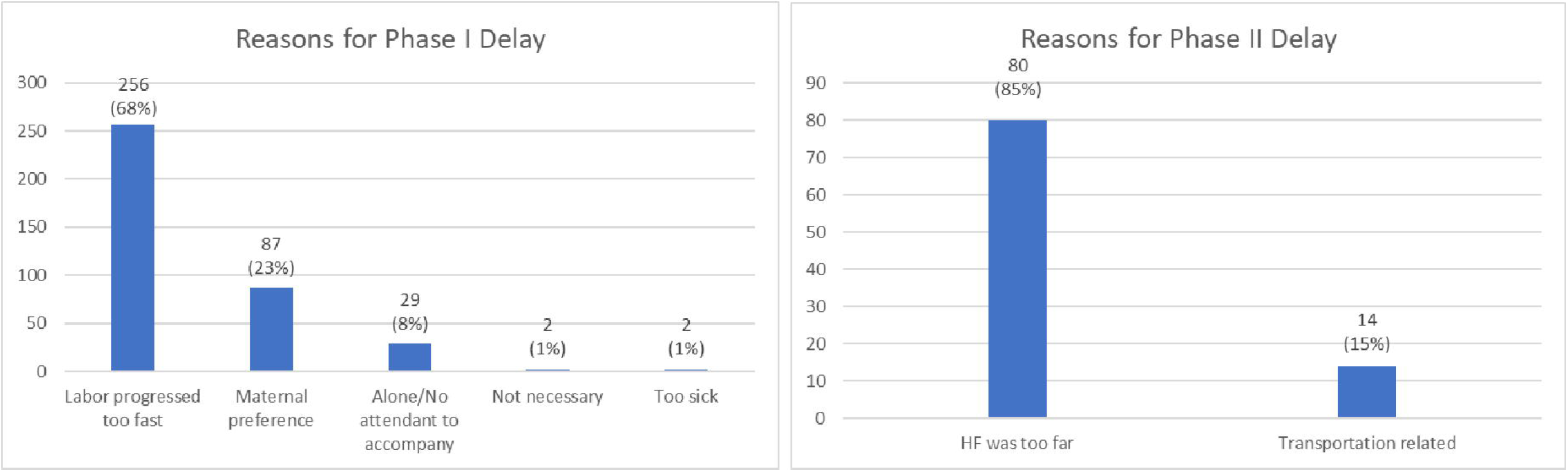
Maternal reasons for Phase I and Phase II delay in the community among women enrolled in the Birhan MCH cohort 2018-2020

### Sensitivity analyses

A total of 173 pregnant women (6.2%) were lost to follow up. Assuming all these women delivered in the community and had no skilled delivery attendance, the point estimate for prevalence of facility birth would decrease from 73.5% to 68.7%, and skilled delivery attendance would decrease from 74.3% to 69.4%. With the assumption that all these women delivered in health facilities, the prevalence would increase from 73.5% to 75.2% for facility birth and 74.3% to 75.9% for skilled delivery attendance. Therefore, the possible range of prevalence for facility births was 68.7 to 75.2%, and for skilled delivery attendance was 69.4 to 75.9%.

## Discussion

This study provided estimates of health facility delivery which is a key component to track progress towards achieving SDG 3.1 to reduce maternal mortality. We identified that 73.6% of study participants gave birth in a health facility. Furthermore, several individual, social and economic determinants, including number of ANC visits, level of education and wealth index, influenced where women gave birth and are discussed below in greater detail.

We found a higher prevalence of facility births compared to national surveys in Ethiopia. The EDHS 2019 reported 54.2% prevalence of facility births in the Amhara region and 47.5% facility births nationwide (16). Surveys like the EDHS are based on recall of events over the five years preceding the survey and may have limited ability to present timely estimates of coverage. Estimates from other studies conducted at the same time vary, with those from the Amhara region reporting facility birth prevalence in the range of 45% to 62% (24,34–38). Over the past decade, there has been a 200% increase in facility delivery coverage from 10.5% in 2011 to 52.5% in 2019 (15,16,39), largely attributed to the work of the Health Development Army (HDA), prohibition of the use of traditional birth attendants (TBAs), increase in the number of staffed health centers, and improved transport infrastructure (40–42). Our estimates of facility-based delivery and skilled birth attendance align with the increasing trend in coverage observed through the past years.

Our study explored factors under the broad classification of socio-cultural (age, ethnicity, religion, education), economic accessibility (wealth index), physical accessibility (place of residence, distance to health facility) and perceived benefit/need of skilled attendance (number of ANC visits, parity, day of birth) based on the conceptual framework proposed by Sabine Gabrysch et al (32). We identified that women who lack any formal education, come from poorest or poorer wealth quintile, reside over an hour walking distance from health facility and had no ANC follow up during pregnancy are less likely to deliver in a health facility. These findings are comparable to those from other studies in low-income countries where they were shown to be important factors affecting where women deliver (17,43–46).

Maternal formal education influences health seeking behavior and health related decisions in different pathways. Formal education curricula provide health, hygiene, nutrition and care-seeking messages and enrich women’s health knowledge base (47). Women with more education tend to seek additional sources of information from media and understand educational messages better, are more receptive to health updates, more influential in decision making processes, and in turn, are able to make better health decisions (45,48–53). Similarly, wealth is an important determinant of facility delivery as women and families with higher resources are likely to have better awareness, ability to pay for and access information and services (49,52–54). This finding may be an important marker of inequality that highlights the recent gains in health facility delivery have left behind certain subgroups of the population who require targeted messages, interventions and continued investment on economic welfare.

Women with higher parity are more inclined to deliver at home, which could be due to prior favorable child-birth experiences and lower felt need for assistance (55). Educational and other interventional programs can be specifically designed to target these subgroups to improve awareness regarding the significance of skilled delivery care. Similar to other studies in low income countries, we show that having one or more ANC visits was associated with better uptake of facility-based services, likely due to the benefit of familiarizing women with the health system, antenatal counselling and birth preparedness packages (20,56). Prior studies have shown that attending the number of ANC visits recommended by WHO provides frequent contacts with health facility and opportunities to improve trust with providers, and deliver more counselling and key interventions (57).

Evidence from previous studies that included distance as a determinant have shown less use of health facility delivery care with increasing distance from health facilities (58–60) and link to adverse health outcomes (61–63). Distance to health facilities presents a dual obstacle to women by discouraging them from seeking initial care and by presenting challenges to reaching facilities after they have made the decision to seek care (64). Dotse-Gborgbortsi et al have shown that a kilometer increase in distance reduced likelihood of health facility delivery by 6.7% (59). In this study, we have quantified distance in terms of travel time that encompasses not only linear distance, but also includes challenges related to modes of transportation to better reflect the effect on the decision-making process. Measurement and analysis of travel times is a useful way to better identify underserved areas and those at increased need of access to services to ensure equitable distribution of services.

The most cited reason for not delivering in a health facility was linked to Phase I in the delay model with participants reporting a delay in deciding to seek care. The majority of the women in our cohort had the perception that their labor progressed too fast and felt they did not have enough time to make decisions. This could be due to poor knowledge and awareness of the signs and progress of labor. Many women also expressed their personal preference to have deliveries at home, which may relate to a lack of awareness of the benefits of skilled delivery attendance, prior self or family experiences with pregnancy and delivery, as well as perception of low risk for the index pregnancy (32). With the recognition that every pregnancy is at risk, there is a need to educate women on the importance of a birth plan during ANC that outlines how to access the closest health facility that has the capacity to perform deliveries and handle complications.

Phase II in the delay model, delay in reaching the health facility, was also a common reason mentioned by women in our study for not delivering in a health facility. Over the past years, the Ethiopian government has made investments in improving physical accessibility to care in health facilities with the establishment of Maternity Waiting Homes (MWH). While this initiative offers an inexpensive method of improving access to health services, studies have shown that there is poor uptake of these services due to lack of awareness, lack of transportation to the MWHs, shortage of food and basic supplies in these MWHs, and lack of child care for younger children at home (65). This calls for intensified community awareness activities about the importance and utilization of MWHs as well as their incorporation into individual birth preparedness and readiness plans, especially for those women with significant challenges related to access. Further assessment of the condition of MWHs and services needs to be made to improve the situation and cater to the needs of mothers and families. Additionally, locally sourced solutions need to be sought for women and families with most severe challenges.

### Strengths and Limitations

A strength of this study is its contribution of longitudinal data that includes community and facility-based ascertainment of where and when pregnancy outcomes occur, in a large and representative sample population. Additionally, we measure and describe associations in terms of relative risk which is a more accurate estimation of the relationship between the covariates and a common outcome. Our analysis of influence of determinants on the study outcome highlight key areas and subgroups of the population to target for further interventions. This may serve as a useful model that can be used in different settings to understand the true burden of the identified challenges.

As a limitation, it is worth noting the effect of study related community surveillance activities on encouraging uptake of facility-based services during pregnancy and delivery. To the extent possible, the study implemented an observational design and the proportion of deliveries in health facilities have remained fairly constant throughout the study. The presence of any Hawthorne effect is anticipated to be insignificant. Approximately 6% of pregnancies were lost to follow up, but this was not expected to alter findings as sensitivity analyses demonstrated limited variation in the outcome estimate based on assumed scenarios of prevalence from missing data. Additional assessment of decision-making roles in the household may have been helpful to understand the role of participant women and specific family members in key decisions that prevent delays in seeking and receiving skilled care.

## Conclusion

The coverage of health facility delivery is increasing in Ethiopia, but falls below the national and global targets. There are significant inequalities in access and utilization of skilled care during childbirth. This study has identified and measured the multidimensional determinants to highlight gaps and areas of focus. Increased investment in formal education, ANC attendance and access to care, among others, are needed to ensure equitable access and utilization. The design of interventions should focus on those with otherwise unfavorable determinants such as women from the poor wealth quintiles, those with high parity and those with long travel times to health facilities. Efforts to maximize the use of existing interventions should include education of mothers on the signs and progress of labor during ANC visits, as well as making facility-based delivery services accessible and usable by mothers with unfavorable predictors.

## Data Availability

Data use is governed by the Birhan Data Access Committee (DAC) and follows Birhan data sharing policy. All researchers who wish to access Birhan data can complete a Birhan data request form and submit it for decision by the Birhan DAC.

## Acknowledgements

We would like to thank all mothers and families who participated in the study and the community of the Birhan field site. We thank the data collectors, supervisors and the HaSET team for their contributions.

## Supporting information

S1 File: Multivariate poisson regression for determinants of health facility delivery

